# Serum Procalcitonin as an Early Predictor of Postoperative Complications Following Liver Resection: A Prospective Observational Study

**DOI:** 10.1101/2025.11.20.25340683

**Authors:** Mohammad Saydul Aman, Md. Shawkat Ali, Bishow Raj Khadka, Bineet Kumar Yadav, Bidhan Chandra Das, Md Foyjul Islam

## Abstract

Postoperative complications (POC) following liver resection (LR) can significantly affect patient outcomes, and early predictors are crucial for timely intervention, particularly in resource-limited settings such as Bangladesh. In this prospective observational study conducted from August 2021 to July 2022, 42 patients undergoing LR were enrolled and divided into two groups based on the presence (n = 22) or absence (n = 20) of POC. Serum procalcitonin (PCT), white blood cell count (WBC), and C-reactive protein (CRP) were measured on the first, third, and seventh postoperative days (POD). Mean serum PCT levels were significantly higher in patients with POC on all PODs (first POD: 2.508 vs. 0.898 µg/L, p = 0.011; third POD: 0.890 vs. 0.314 µg/L, p = 0.002; seventh POD: 0.611 vs. 0.181 µg/L, p = 0.005), whereas WBC and CRP did not differ between groups. The area under the receiver operating characteristic curve (AUC) for serum PCT was 0.798, 0.797, and 0.769 on the first, third, and seventh POD, respectively, indicating that first POD PCT is a reliable early predictor of POC. A cutoff value of 1.100 µg/L achieved 86.4% sensitivity and 70% specificity. These findings suggest that serum PCT on the first POD is a promising biomarker for early detection of postoperative complications after liver resection.

## Introduction

Liver resection (LR) is regarded as the gold-standard surgical approach for various liver lesions. Despite being a highly invasive procedure, it has become safer in recent years. Appropriate patient selection, advances in surgical and anesthetic techniques, the use of minimally invasive surgery, and improvements in perioperative care have expanded the indications for elective LR. In the last two decades, these developments have reduced post-operative mortality from more than 10% to less than 5% [1–3]. However, post-operative morbidity remains high, at around 20-45% in various studies [3–5].

Generally, early detection of postoperative complications (POC) after elective LR is a challenge for liver surgeons. Various parameters such as white blood cell (WBC), C-reactive protein (CRP), interleukin-6 (IL-6), international normalized ratio (INR), serum albumin, serum bilirubin, serum creatinine, serum transaminases, etc., are commonly used for early prediction with variable results [6–10]. Most of these either exhibited low sensitivity or low specificity for morbidity and mortality [3]. The commonly used CRP usually peaks on the second or third post-operative day (POD) and reduces gradually afterwards [11]. Most of the other markers are not useful prior to the fifth POD. Moreover, multiple studies have shown that peri-operative steroids can cause a significant reduction of some markers (CRP, IL-6) in the early post-operative period [11,12]. As a result, it has become difficult to use these markers for prediction.

Procalcitonin (PCT), the 116-amino-acid precursor of calcitonin, is generally produced by thyroid C-cells in healthy people [13]. Various types of stimuli, such as surgery and trauma, alter this typical source of PCT synthesis. Many extra-thyroid tissues (e.g., liver, testis, lung, kidney, etc.) highly express the calcitonin gene CALC-1 [14]. Also, hepatic macrophages have very high expression of PCT-mRNA. Research has proven that acute hepatocyte injury upregulates PCT synthesis in the liver [15,16]. So, after surgery, the PCT concentration may increase 100-fold to 10,000-fold from a baseline of 0.5 µg/L. Steroid administration has no effect on its production. Serum PCT has a 25-30 hour half-life and can reach peak levels within 18-24 hours post-surgery [10,16,17]. A drop in PCT level (compared to the previous day’s value) indicates that the ongoing treatment is effective, and the prognosis is favorable. On the other hand, a persistently raised value over time is associated with a worse prognosis and may indicate an impending complication.

Safe liver resection is paramount for a safe outcome. In low-income countries like Bangladesh, where resources are limited and data regarding elective LR is scarce, an early predictor is essential to ensure a safe outcome. Recently, serum PCT is being measured in the early postoperative period after many major surgeries in resource-rich settings in developed countries as a better prognostic indicator and therapeutic guide [10,11,18]. Some studies suggest that measuring serum PCT between 12 and 48 hours after elective LR could help figure out an adverse outcome, like post-hepatectomy liver failure (PHLF) or death [19,20]. However, there are no such studies on the South Asian cohort that show how serum PCT changes after elective LR. Its impact on clinical practice requires proper evaluation by a prospective study.

This prospective observational study aimed to identify the role of serum PCT levels in the early prediction of POC after elective LR in comparison to common markers (WBC and CRP).

## Materials and methods

### Study design

This prospective observational study was conducted from August 2021 to July 2022 in the Department of Hepatobiliary, Pancreatic, and Liver Transplant Surgery of the Bangladesh Medical University, Dhaka. Patients were enrolled based on the following inclusion criteria: (a) patients undergoing elective LR; (b) age ≥ 18 years; (c) any gender; and (d) Child-Pugh A or B. The exclusion criteria were: (1) LR following trauma; (2) Child-Pugh C; (3) any unplanned LR; (4) patients without written informed consent; and (5) patients with incomplete post-operative data.

### Data collection, surgical procedure, operational definitions, and clinical management

Considering the enrolment criteria, forty-two consecutive patients were enrolled in this study. All of them were evaluated preoperatively through proper history-taking, physical examination, laboratory investigations, and imaging studies. Before every surgery, a clinical discussion meeting was conducted to ensure that LR was well planned. The general condition of all patients was optimized to improve their performance status [21]. All patients were evaluated pre-operatively by anesthesiologists using the American Society of Anesthesiologists score [22].

All patients underwent LR under general anesthesia with tracheal intubation. They were operated on through an open approach (right subcostal or reverse L incision). After a thorough assessment of the peritoneal cavity, attention was given to hepatic inflow and outflow control. Parenchymal transection was done either by the clamp-crush technique or by using an ultrasonic dissector, depending on the surgeon’s choice. If required, concomitant procedures were also performed based on indications (e.g., biliary bypass for recurrent hepatolithiasis, radical lymphadenectomy for gall bladder malignancy, etc.). Special attention was given to proper hemostasis and the prevention of bile leakage. All the resected specimens were sent for histopathology. “The Brisbane 2000 terminology of liver anatomy and resections” was used to define all procedures, e.g., name, type, extent, order, etc., for uniform documentation [23,24]. Major resection was defined as a resection of three or more contiguous or non-contiguous liver segments [2,7]. After recovery, all patients were kept in the post-operative ward overnight and shifted to their respective wards the next morning. No patients received perioperative steroids.

Within the next 30 days, if any POC happened, it was noted and dealt with according to standard hospital protocol. Standard operational definitions of the International Study Group of Liver Surgery were used for defining PHLF [25], post-hepatectomy hemorrhage (PHH) [26], and bile leak [27]. Other complications were also defined according to the standard protocol.

Following hospital discharge, all patients were seen in the outpatient department within the next 30 days. All patient-identifying data was kept confidential and encoded before being entered into a database. The highest levels of confidentiality and ethical standards were maintained during data storage and analysis.

### PCT measurement

Serum samples were collected in the mornings of the 1st, 3rd, and 7th POD from a peripheral vein and sent for WBC count, CRP, and PCT measurement. The PCT assay was done in the Department of Biochemistry & Molecular Biology of the same hospital and the Siemens Atellica IM BRAHMS Procalcitonin (PCT) analyzer (reference 11202699) was used with a measuring interval of 0.03–50.00 µg/L.

WBC count and CRP concentration were generally used as predictive markers after surgery in our hospital. By contrast, because no clear criteria for the PCT concentration were available during the study period, the PCT level did not affect the clinical treatment of the patients. Based on the patient’s clinical status, if necessary, any other hematological, biochemical, or imaging investigations were done and repeated.

### Grouping

After surgery, all patients were divided into two groups depending on the absence or presence of POC. Group A included patient with no POC after surgery. Group B included patients with POC after surgery. We examined the relationship between predictive markers (WBC count, CRP, and PCT) and POC.

### Statistical analysis

Statistical analyses were carried out using the Statistical Package for Social Sciences (SPSS) version 25.0 by SPSS Inc. (IBM), New York, USA. Categorical (qualitative) variables were presented as frequencies and percentages. The chi-square (χ^2^) test, or Fisher’s exact test, was performed to check for any association between groups, where appropriate. Continuous (quantitative) variables were presented as mean ± standard deviation (SD). The differences between groups were compared using the independent sample t-test. The area under the receiver operating characteristics (ROC) curve (AUC) was calculated to identify a cutoff value for serum PCT. The statistical tests were conducted with a 95% confidence interval (CI), and p < 0.05 was considered statistically significant.

## Results

A total of 42 patients undergoing liver resection were included in the analysis. Among them, 20 patients developed postoperative complications (Group A), while 22 did not (Group B) Minor resections were most common (78.6%), with similar distribution between patients who developed complications (Group A: 80.0%) and those who did not (Group B: 77.3%) (p = 0.830). Anatomical resections predominated overall (71.4%), with no significant difference between groups (p = 0.379). The most frequent operations were hepatic bisegmentectomy (38.1%) and left lateral sectionectomy (33.3%), but complication rates did not differ across procedures (p = 0.650). Resection order (first-, second-, or third-order) was comparable between groups (p = 0.810). Use of the Pringle maneuver (33.3% overall) and transection technique (clamp-crush vs. ultrasonic dissector) also showed no significant association with postoperative complications (p = 0.662 and p = 0.746, respectively). [Table1]

**Table 1.** Distribution of study participant by Operative procedures (N = 42)

| Variable | Total n (%) | Group A<br>(n = 20) | Group B<br>(n = 22) | p-value* |
| --- | --- | --- | --- | --- |
| **Extent of resection** |  |  |  |  |
| Minor (<3 segments) | 33 (78.6) | 16 (80.0) | 17 (77.3) | **0.830** |
| Major ($\geq 3$ segments) | 9 (21.4) | 4 (20.0) | 5 (22.7) | |
| Type of resection |  |  |  |  |
| Anatomical | 30 (71.4) | 13 (65.0) | 17 (77.3) | 0.379 |
| Non-anatomical | 12 (28.6) | 7 (35.0) | 5 (22.7) |  |
| Name of operation |  |  |  |  |
| Hepatic bisegmentectomy<sup>a</sup> | 16 (38.1) | 8 (40.0) | 8 (36.4) | 0.650** |
| Left lateral sectionectomy((i.e. bisegmentectomy 2, 3)) | 14 (33.3) | 6 (30.0) | 8 (36.4) |  |
| Left hepatectomy | 6 (14.3) | 3 (15.0) | 3 (13.6) |  |
| Right hepatectomy | 2 (4.8) | 1 (5.0) | 1 (4.5) |  |
| Right posterior sectionectomy | 2 (4.8) | 2 (10.0) | 0 (0.0) |  |
| Segmentectomy 6 | 1 (2.4) | 0 (0.0) | 1 (4.5) |  |
| Left lateral sectionectomy + segmentectomy 6 | 1 (2.4) | 0 (0.0) | 1 (4.5) |  |
| Order of resection |  |  |  |  |
| First-order | 8 (19.0) | 4 (20.0) | 4 (18.2) | 0.810 |
| Second-order | 16 (38.1) | 8 (40.0) | 8 (36.4) |  |
| Third-order | 17 (40.5) | 8 (40.0) | 9 (40.9) |  |
| Extended | 1 (2.4) | 0 (0.0) | 1 (4.5) |  |
| Inflow occlusion (Pringle) |  |  |  |  |
| No | 28 (66.7) | 14 (70.0) | 14 (63.6) | 0.662 |
| Yes | 14 (33.3) | 6 (30.0) | 8 (36.4) |  |
| Transection technique |  |  |  |  |
| Clamp-crush | 20 (47.6) | 9 (45.0) | 11 (50.0) | 0.746 |
| Ultrasonic dissector | 22 (52.4) | 11 (55.0) | 11 (50.0) |  |
| Concomitant intra-abdominal procedure |  |  |  |  |
| No | 17 (40.5) | 13 (65.0) | 4 (18.2) | 0.002 |
| Yes | 25 (59.5) | 7 (35.0) | 18 (81.8) |  |

Data are presented as n (%). <sup>a</sup> Hepatic bisegmentectomy, which included Bisegmentectomy 4b, 5, Bisegmentectomy 5, 6 etc. \* Chi-square test \*\* Fisher exact test.

In both groups, patients’ mean age, mean body mass index, performance status, ASA score, and Child-Pugh score were comparable. Regarding gender, male patients were more associated with POC than female patients in our study (15 vs. 7, p < 0.05). Except for diabetes, other comorbidities had no statistically significant association with POC. The mean MELD score was statistically significant between both groups (6.70 ± 1.30 vs. 8.82 ± 3.80, p < 0.05). However, other pre-operative hematological and biochemical functions were comparable between the two groups. [Table 2]

**Table 1.** Comparison of baseline patient characteristics between two groups (all patients = 42).

| Variables | Group A<br>(n = 20) |  | Group B<br>(n = 22) |  | P value |
| --- | --- | --- | --- | --- | --- |
| Age, years, mean ± SD | 46.95 ± 13.05 | | 47.05 ± 15.15 | | 0.983 |
| Sex of the participants, n (%) |  |  |  |  | **0.013** |
| • Male | 6 | (30) | 15 | (68.2) |  |
| • Female | 14 | (70) | 7 | (31.8) |  |
| BMI, kg/m<sup>2</sup>, mean ± SD | 21.32 ± 4.09 | | 22.02 ± 3.60 | | 0.806 |
| WHO performance status, n (%) |  |  |  |  | 0.297 |
| • 0 | 16 | (80) | 14 | (63.6) |  |
| • 1 | 4 | (20) | 5 | (22.7) |  |
| • 2 | 0 | (0) | 3 | (13.6) |  |
| ASA score, n (%) |  |  |  |  | 0.123 |
| • I | 8 | (40) | 14 | (63.6) |  |
| • II | 11 | (55) | 5 | (22.7) |  |
| • III | 1 | (5) | 3 | (13.6) |  |
| Child-Pugh score, n (%) |  |  |  |  | 1.000 |
| • A | 19 | (95) | 20 | (90.9) |  |
| • B | 1 | (5) | 2 | (9.1) |  |
| MELD score, mean ± SD | 6.70 ± 1.30 | | 8.82 ± 3.80 | | **0.021** |
| Co-morbidities, n (%) | 16 | (80) | 14 | (63.5) | 0.241 |
| • Hypertension | 8 | (40) | 5 | (22.7) | 0.227 |
| • Diabetes | 9 | (45) | 3 | (13.6) | **0.025** |
| • Previous upper abdominal surgery | 5 | (25) | 5 | (22.7) | 1.000 |
| • Respiratory (Asthma/COPD) | 1 | (5) | 5 | (22.7) | 0.187 |
| • Cardiac (IHD) | 2 | (10) | 2 | (9.1) | 1.000 |

| Variables | Group A |  | Group B |  | P value |
| --- | --- | --- | --- | --- | --- |
|  | (n = 20) |  | (n = 22) |  |  |
| Renal (CKD) | 1 | (5) | 2 | (9.1) | 1.000 |
| Baseline hematological parameters, mean ± SD |  |  |  |  |  |
| Hemoglobin, g/dL | 11.84 ± 1.31 |  | 12.64 ± 2.11 |  | 0.144 |
| Hematocrit, % | 36.67 ± 3.51 |  | 38.17 ± 5.63 |  | 0.302 |
| Red blood cells, x10<sup>12</sup>/L | 4.52 ± 0.51 |  | 4.52 ± 0.68 |  | 1.000 |
| WBC, x10<sup>9</sup>/L | 8.36 ± 2.03 |  | 7.94 ± 2.17 |  | 0.521 |
| Platelet, x10<sup>9</sup>/L | 318.65 ± 89.15 |  | 261.32 ± 107.08 |  | 0.068 |
| Baseline biochemical parameters, mean ± SD |  |  |  |  |  |
| Serum total bilirubin, mg/dL | 0.56 ± 0.25 |  | 1.57 ± 3.35 |  | 0.171 |
| Serum albumin, g/L | 40.90 ± 4.44 |  | 41.21 ± 6.43 |  | 0.856 |
| INR | 0.97 ± 0.14 |  | 1.02 ± 0.17 |  | 0.279 |
| ALT, U/L | 31.80 ± 22.71 |  | 43.45 ± 21.60 |  | 0.096 |
| Serum creatinine, mg/dL | 0.86 ± 0.19 |  | 1.00 ± 0.38 |  | 0.145 |
| eGFR, ml/min/1.73 m<sup>2</sup> | 88.55 ± 23.99 |  | 92.18 ± 32.53 |  | 0.685 |

Data are presented as n (%) or mean ± standard deviation. SD, standard deviation; BMI, body mass index; WHO, World Health Organisation; ASA, American Society of Anesthesiologists; MELD, model for end-stage liver disease; COPD, chronic obstructive pulmonary disease; IHD, ischemic heart disease; CKD, chronic kidney disease; WBC, white blood cells; INR, international normalized ratio; ALT, alanine aminotransferase; eGFR, estimated glomerular filtration rate.

In our study, there were 26 benign cases and 16 malignant cases for which elective LR was performed). Among benign cases, the most common indication was bilateral hepatolithiasis and symptomatic giant hepatic hemangioma (seven cases each). All patients with bilateral hepatolithiasis were faced with a POC after surgery (p < 0.05). Among the malignant cases, the most common indication of LR was hepatocellular carcinoma and gall bladder mass (eight cases each). [Table3]

**Table 2:**
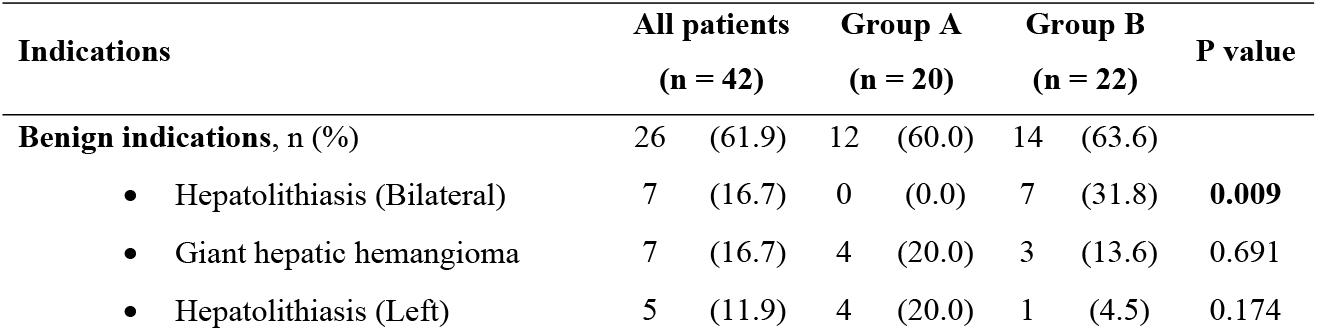
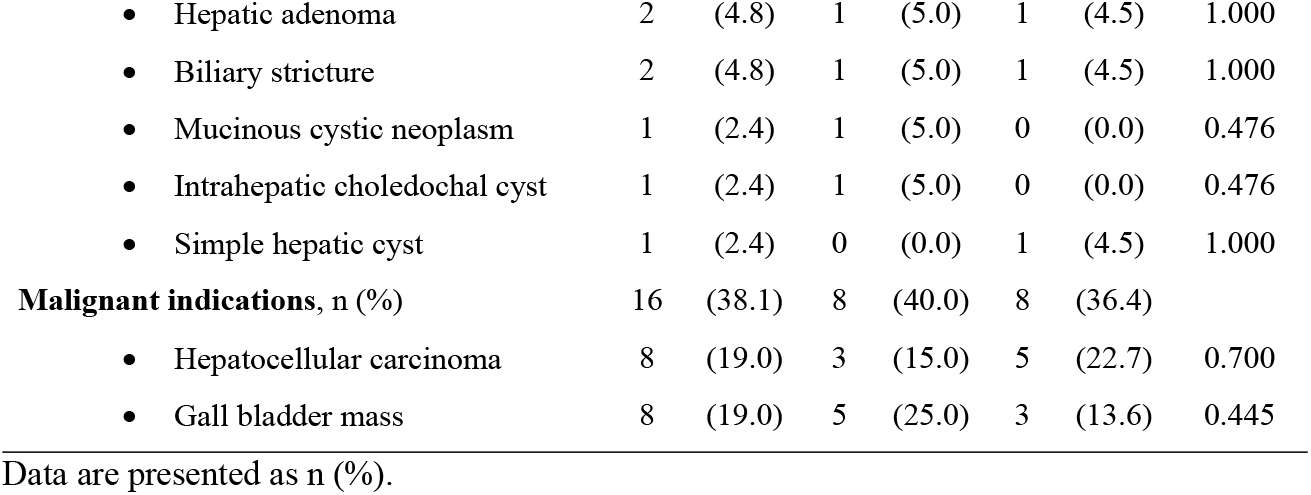
Indications of Liver Resection.

| Indications | All patients<br>(n = 42) |  | Group A<br>(n = 20) |  | Group B<br>(n = 22) |  | P value |
| --- | --- | --- | --- | --- | --- | --- | --- |
| **Benign indications, n (%)** | 26 | (61.9) | 12 | (60.0) | 14 | (63.6) |  |
| • Hepatolithiasis (Bilateral) | 7 | (16.7) | 0 | (0.0) | 7 | (31.8) | **0.009** |
| • Giant hepatic hemangioma | 7 | (16.7) | 4 | (20.0) | 3 | (13.6) | 0.691 |
| • Hepatolithiasis (Left) | 5 | (11.9) | 4 | (20.0) | 1 | (4.5) | 0.174 |
| • Hepatic adenoma | 2 | (4.8) | 1 | (5.0) | 1 | (4.5) | 1.000 |
| • Biliary stricture | 2 | (4.8) | 1 | (5.0) | 1 | (4.5) | 1.000 |
| • Mucinous cystic neoplasm | 1 | (2.4) | 1 | (5.0) | 0 | (0.0) | 0.476 |
| • Intrahepatic choledochal cyst | 1 | (2.4) | 1 | (5.0) | 0 | (0.0) | 0.476 |
| • Simple hepatic cyst | 1 | (2.4) | 0 | (0.0) | 1 | (4.5) | 1.000 |
| **Malignant indications, n (%)** | 16 | (38.1) | 8 | (40.0) | 8 | (36.4) |  |
| • Hepatocellular carcinoma | 8 | (19.0) | 3 | (15.0) | 5 | (22.7) | 0.700 |
| • Gall bladder mass | 8 | (19.0) | 5 | (25.0) | 3 | (13.6) | 0.445 |

Data are presented as n (%).

It is noteworthy that, Concomitant intra-abdominal procedures were significantly more frequent in Group A than Group B (65.0% vs. 18.2%, p = 0.002). Operative time and use of the Pringle maneuver did not differ significantly between groups (210 ± 86.4 vs. 216 ± 78.2 min, p = 0.810; 30% vs. 36.4%, p = 0.662). Parenchymal transection technique (ultrasonic dissector vs. clamp-crush) was comparable between groups (55% vs. 50%, p = 0.746). Notably, patients with complications had significantly lower mean blood loss compared to those without complications (325 ± 181 mL vs. 475 ± 283 mL, p = 0.046) and shorter postoperative hospital stays (10.2 ± 4.0 vs. 15.9 ± 9.7 days, p = 0.018). [Table 4]

**Table 3:**
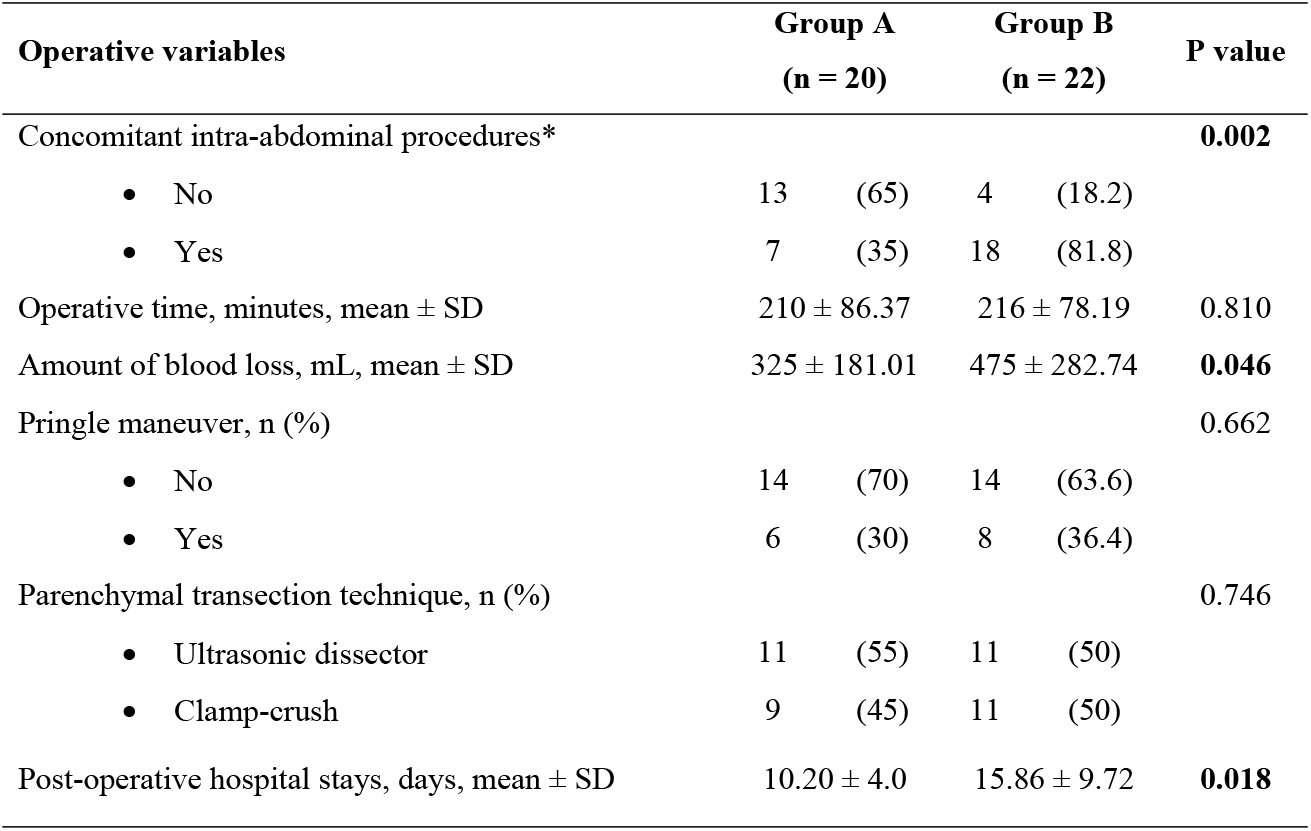
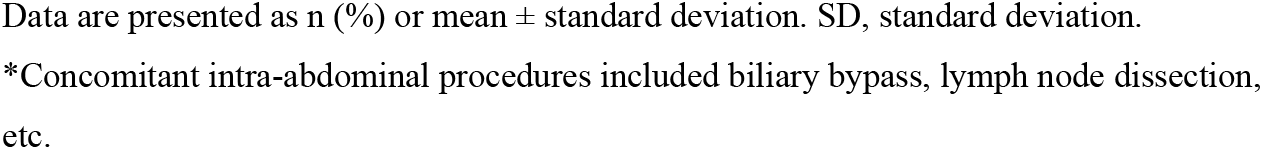
Comparison of operative variables between groups (all patients = 42)

| Operative variables | Group A<br>(n = 20) |  | Group B<br>(n = 22) |  | P value |
| --- | --- | --- | --- | --- | --- |
| Concomitant intra-abdominal procedures* |  |  |  |  | **0.002** |
| • No | 13 | (65) | 4 | (18.2) |  |
| • Yes | 7 | (35) | 18 | (81.8) |  |
| Operative time, minutes, mean ± SD | 210 ± 86.37 |  | 216 ± 78.19 |  | 0.810 |
| Amount of blood loss, mL, mean ± SD | 325 ± 181.01 |  | 475 ± 282.74 |  | **0.046** |
| Pringle maneuver, n (%) |  |  |  |  | 0.662 |
| • No | 14 | (70) | 14 | (63.6) |  |
| • Yes | 6 | (30) | 8 | (36.4) |  |
| Parenchymal transection technique, n (%) |  |  |  |  | 0.746 |
| • Ultrasonic dissector | 11 | (55) | 11 | (50) |  |
| • Clamp-crush | 9 | (45) | 11 | (50) |  |
| Post-operative hospital stays, days, mean ± SD | 10.20 ± 4.0 |  | 15.86 ± 9.72 |  | **0.018** |

Data are presented as n (%) or mean ± standard deviation. SD, standard deviation.
\*Concomitant intra-abdominal procedures included biliary bypass, lymph node dissection, etc.

Among the POC, the most common were PHH grade A (five cases, 11.9%) and PHLF grade B (five cases, 11.9%). Three cases (7.1%) had bile leakage, three cases (7.1%) had wound dehiscence, and two cases (4.8%) had wound infection. [Fig 1] In this one-year LR series, there were no 30-day mortalities.

**Fig 1:**
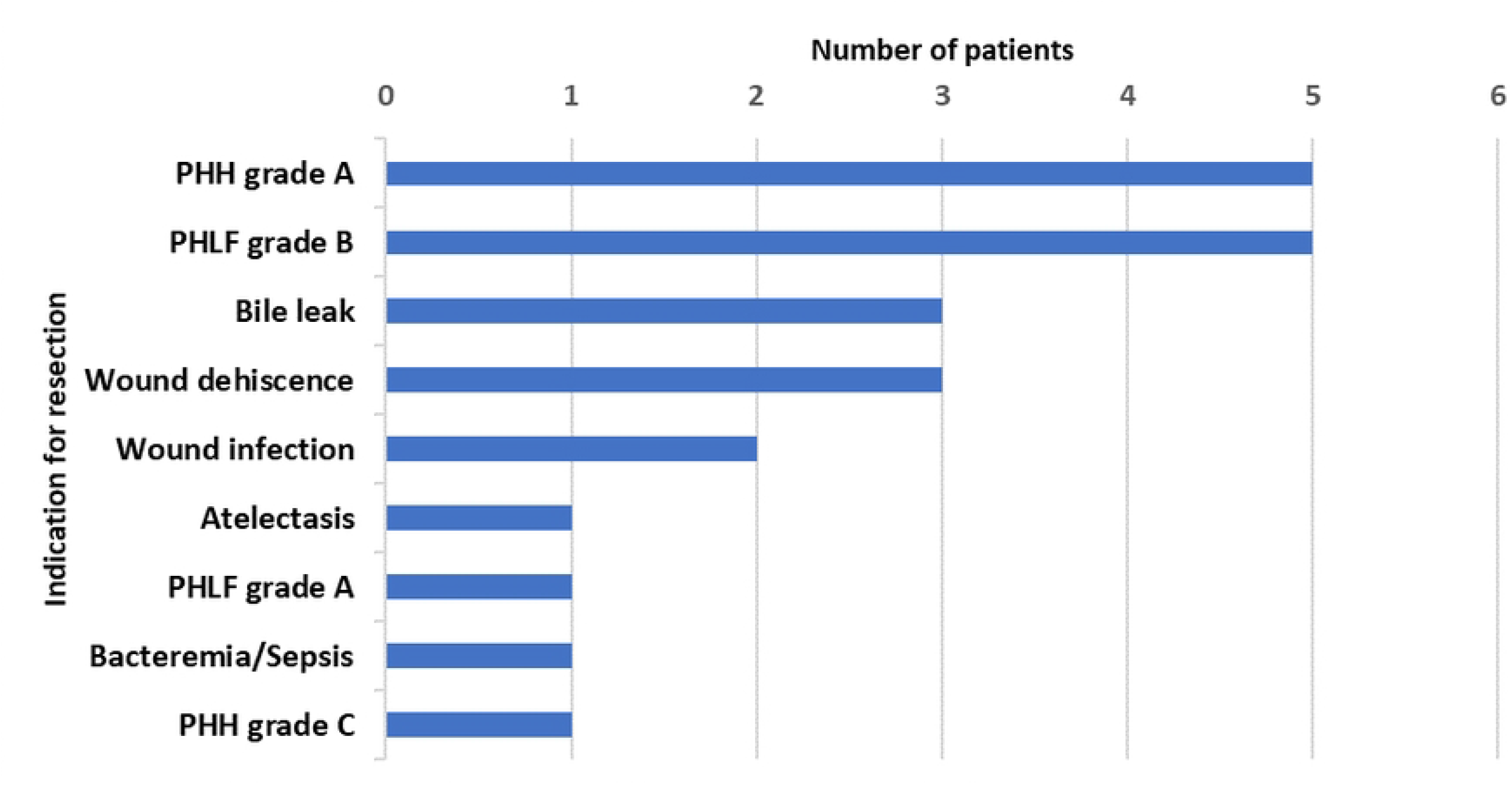
Distribution of study participant by Post-Operative Complications (POC) n (22)

Table 6 shows the comparison of the mean values of WBC, CRP, and serum PCT on POD 1, 3, and 7 between the two groups. The difference in the mean WBC counts (see also Figure 1-A) between the two groups was not statistically significant on all PODs (p = 0.755, 0.799, and 0.118, respectively). Similarly, there were no statistically significant differences in the mean CRP levels (see also Figure 1-B) between the two groups on all PODs (p = 0.617, 0.460, and 0.816, respectively). However, the differences in the mean PCT levels (see also Figure 1-C) between the two groups were statistically significant on POD 1, 3, and 7 (p = 0.011, 0.002, and 0.005, respectively). [Table 6]

**Table 4:**
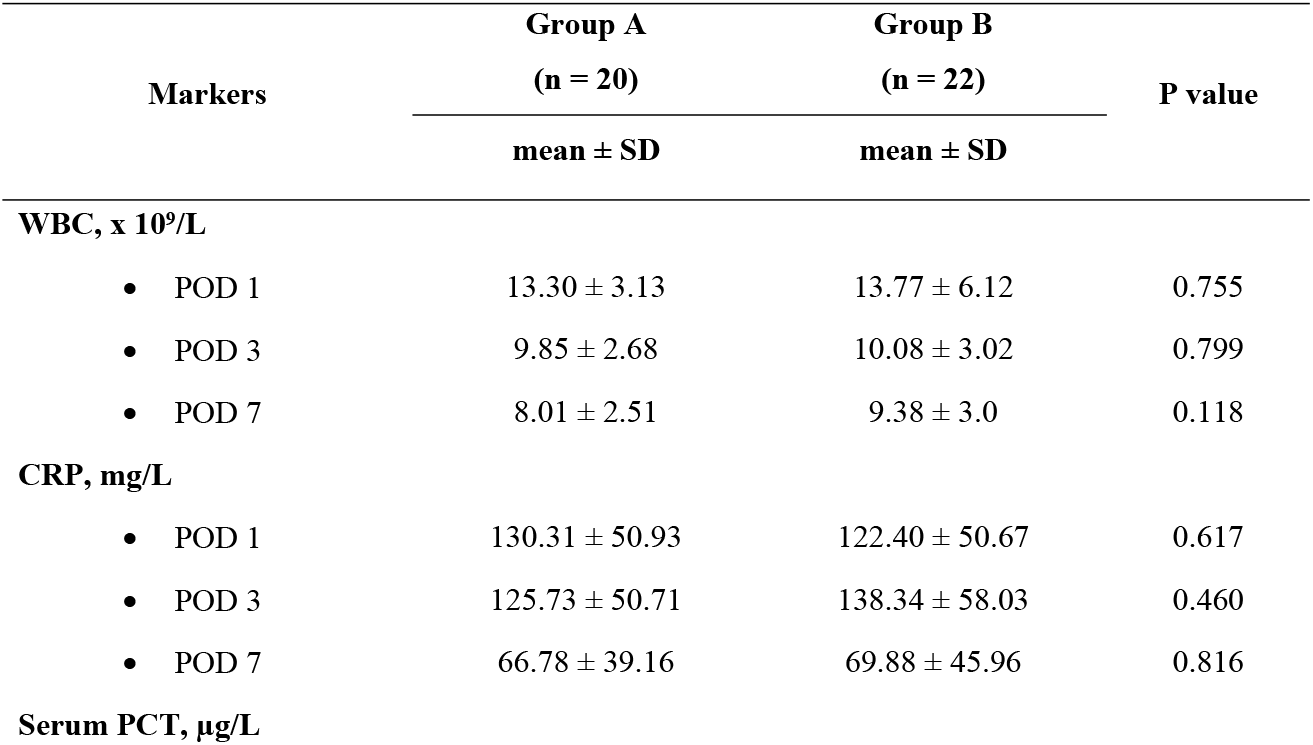
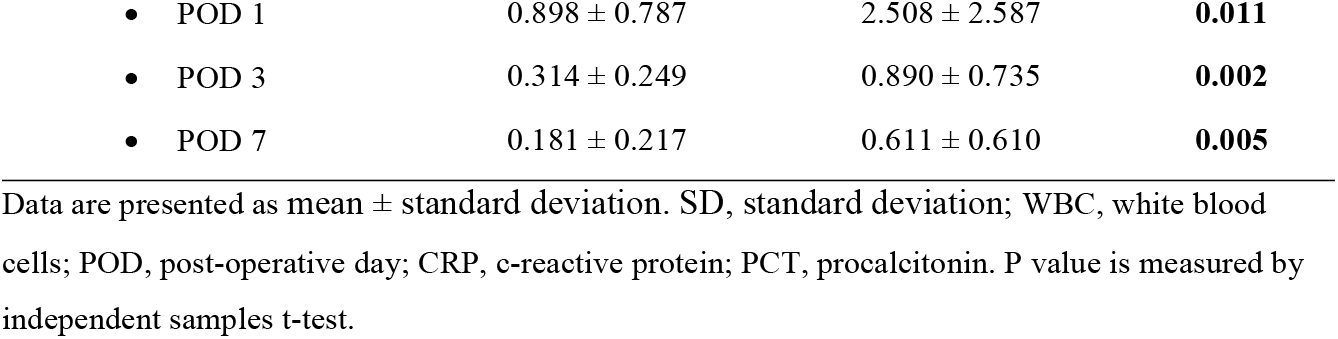
Comparison of the mean values of WBC, CRP, and serum PCT on the post-operative period in between two groups (all patients = 42).

Patients in group B had a higher WBC count and PCT level compared to those in group A throughout the entire postoperative period. [Fig 1-A, 1-C] The differences in mean WBC count observed between the two groups did not reach statistical significance. The mean serum PCT levels exhibited statistically significant differences between the two groups across all PODs (1, 3, and 7). [Table 6] The CRP level was higher on POD 3 and 7 in group B patients compared to group A patient. [Fig 1-B] Nevertheless, these alterations did not reach statistical significance.

Based on these findings, different ROC curves were constructed to assess the predictive power of the POD 1 serum PCT (blue line), POD 3 serum PCT (red line), and POD 7 serum PCT (green line) for the occurrence of POC, using data of all patients in this study. The diagonal line (orange) indicates the reference line. [ Fig 2-D]

**Fig 2:**
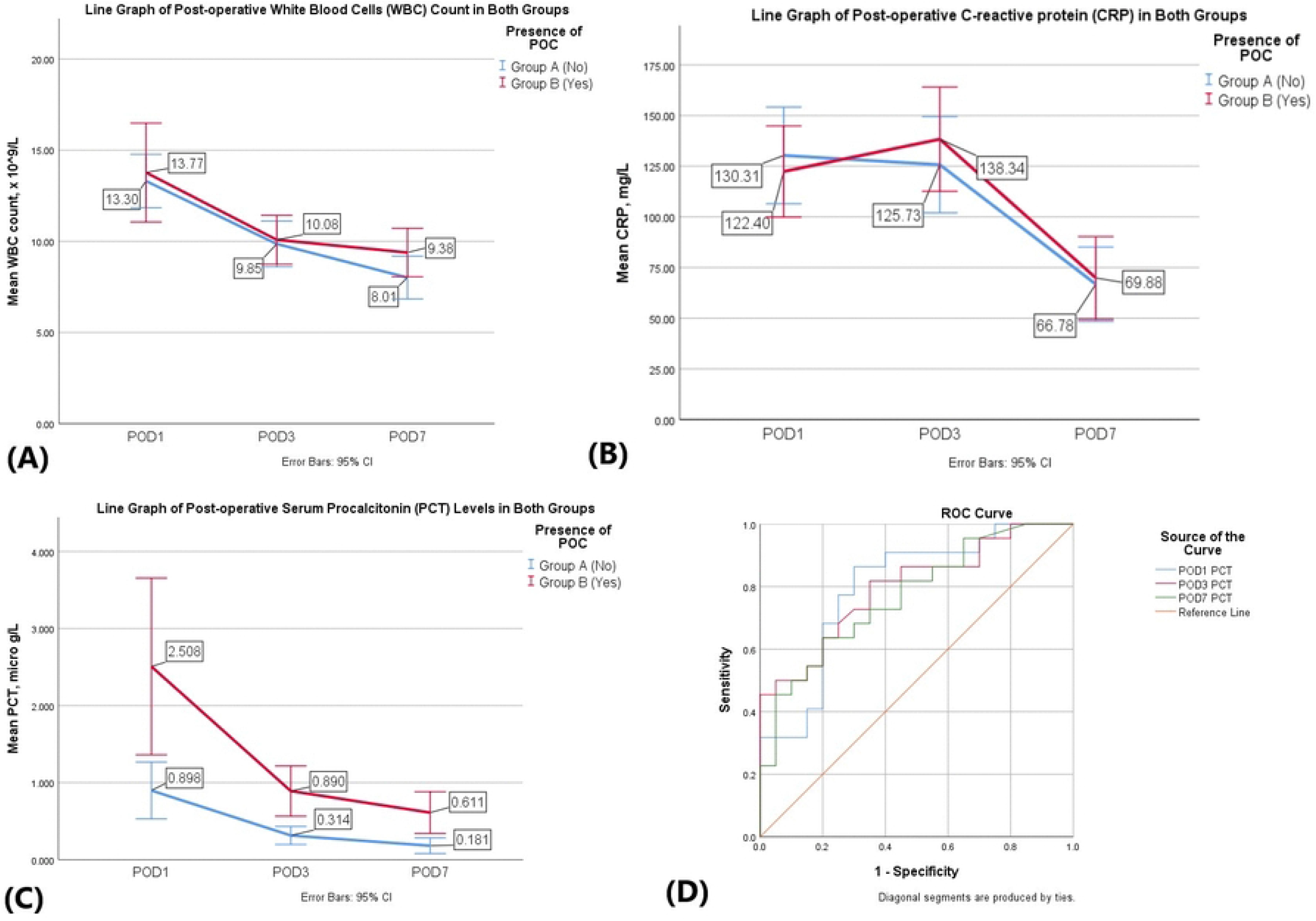
Serial changes in post-operative markers. (A) Comparison of mean WBC count among two groups. (B) Comparison of mean CRP level among two groups. (C) Comparison of mean serum PCT level among two groups. (D) Receiver operating characteristics (ROC) curve of mean serum PCT on post-operative day (POD) 1 (blue line), on POD 3 (red line) and on POD 7 (green line) in predicting post-operative complications (POC) after elective liver resection in all patients. Diagonal line (orange) indicates the reference line.

The AUC for POD 1 serum PCT was 0.798 (95% CI, 0.660, 0.935), POD 3 serum PCT was 0.797 (95% CI, 0.663, 0.930), and for POD 7 serum PCT was 0.769 (95% CI, 0.628, 0.910). Because the AUC for the POD 1 serum PCT was the highest, the best cutoff level was found to be at 1.100 µg/L. This was the point where sensitivity, specificity, positive predictive value (PPV), and negative predictive value (NPV) were 86.4%, 70.0%, 74.2%, and 83.7%, respectively. In order to find an early predictor of POC after elective LR, the POD 1 serum PCT level of 1.100 µg/L was selected as the best possible predictor. [Table 7]

**Table 5:** Area under the curve (AUC) based on the mean serum PCT levels measured on POD 1, 3 and 7.

| Test Result Variables | Area (AUC) | Asymptotic 95% CI | | Cutoff value, $\mu\text{g/L}$ | Sensitivity, % | Specificity, % | PPV, % | NPV, % |
| --- | --- | --- | --- | --- | --- | --- | --- | --- |
|  |  | Lower Bound | Upper Bound |  |  |  |  |  |
| POD 1 PCT | 0.798 | 0.660 | 0.935 | 1.100 | 86.4 | 70.0 | 74.2 | 83.7 |
| POD 3 PCT | 0.797 | 0.663 | 0.930 | 0.405 | 72.7 | 70.0 | 70.8 | 71.9 |
| POD 7 PCT | 0.769 | 0.628 | 0.910 | 0.160 | 72.7 | 65.0 | 67.5 | 70.4 |

AUC, area under the curve; POD, post-operative day; PCT, procalcitonin; CI, confidence interval; PPV, positive predictive value; NPV, negative predictive value.

## Discussion

Liver resection (LR) remains a technically demanding procedure, and although perioperative mortality has declined globally, postoperative morbidity continues to be a major clinical challenge[3]. Evidence from high-income countries shows substantial improvements in perioperative care; however, the applicability of these findings to low-resource settings such as Bangladesh has been insufficiently explored. This prospective study contributes to this gap by evaluating postoperative outcomes following elective LR and, importantly, assessing the utility of serum procalcitonin (PCT) as an early biomarker for postoperative complications (POC).

The present study demonstrates that serum PCT is a more reliable early predictor of POC than conventional inflammatory markers, such as WBC count and CRP. PCT levels peaked within the first postoperative day (POD), consistent with findings from Kretzschmar et al., reflecting its rapid induction following hepatocellular injury [17]. A PCT threshold of 1.100 µg/L on POD 1 showed strong discriminatory ability (AUC 0.798; 95% CI 0.660–0.935), with high sensitivity (86.4%) and acceptable specificity (70%). In contrast, WBC count demonstrated nonspecific postoperative fluctuations, and CRP exhibited its characteristic delayed rise, limiting their value for early risk stratification. These findings reaffirm earlier reports indicating the superior early predictive performance of PCT in major abdominal surgery [10,11].

Our results align closely with those of Li et al., who observed that PCT ≥ 1.000 µg/L at 12 hours post-LR in hepatocellular carcinoma patients was strongly associated with PHLF, sepsis, and mortality[20]. The similarity between these thresholds supports the biological validity of PCT as an inflammatory stress marker after LR. However, the broader range of underlying pathologies included in our study enhances the generalizability of these findings to diverse surgical indications.

PCT is usually regarded as a marker of bacterial infection or sepsis. But it also rises as a pro-inflammatory cytokine after major surgery, such as liver resection [10,11,17]. According to Aoki et al. [19], the peak serum PCT within the first two days of surgery can correctly predict Clavien-Dindo complications grade II and above. They set a cutoff value of 0.35 µg/L (AUC, 0.860), with a sensitivity and specificity rate of 80% and 83%, respectively. However, that cutoff value was below the normal baseline level of serum PCT 0.5 µg/L for any clinical application. Using the serum PCT as a predictor of POC necessitates a value greater than 0.5 µg/L. In that sense, it is hard to utilize “0.35 µg/L” as a benchmark for future reference. In our study, we calculated a new cutoff value for serum PCT of 1.100 µg/L on the first POD after elective LR. It has a higher sensitivity of 86.4%. Interestingly, Aoki et al. [19] did not mention patients with Clavien-Dindo grade I complications. Rather, they added those patients to their non-complication group for study purposes. In our study, we added all patients who developed complications into one group (Group B) for comparison against the non-complication group (Group A).

We have performed elective LR for 26 benign and 16 malignant diseases. We had no 30-day mortality, though the morbidity rate was slightly on the higher side (52.4%) compared to the literature [2,3]. This relatively high rate of morbidity may be due to the careful identification and documentation provided by both clinical and biochemical examinations performed in the post-operative period at our hospital, so that no minor complications were overlooked.

This study has several limitations. The relatively small sample size and single-center design limit statistical power and external validity. Heterogeneity in surgical indications may have introduced confounding physiological differences in postoperative inflammatory responses. Additionally, preoperative PCT levels were not measured due to cost constraints, which prevents confirmation of postoperative relative change.

Nevertheless, our findings hold important clinical implications. Early PCT measurement may support timely triage, guide diagnostic evaluation, and inform postoperative surveillance strategies, particularly in resource-limited settings where judicious use of hospital resources is essential.

## Conclusion

Serum procalcitonin (PCT) on the first postoperative day (POD) is a reliable early indicator of postoperative complications (POC) following elective liver resection. It demonstrates high sensitivity and moderate specificity, and a cutoff value of 1.100 µg/L on the first POD can predict a wide range of postoperative morbidities in the Bangladeshi patient, irrespective of the underlying liver pathology.

**S1 Data Raw anonymized dataset used in the analysis**

## Data Availability

All of the data of the study was available on manuscript and its supplementary file.

## Notes

### Competing Interest Statement

The authors have declared no competing interest.

### Clinical Trial

NA

### Funding Statement

Yes

### Author Declarations

This study was conducted following approval from the Institutional Review Board of Bangladesh Medical University (Registration No. 3530). Written informed consent was obtained from all participants prior to their inclusion in the study. All procedures were performed in accordance with relevant guidelines and regulations, and participant confidentiality was strictly maintained.

